# Exploring the relationship between cultural and structural workforce issues and retention of nurses in general practice (GenRet): A qualitative interview study

**DOI:** 10.1101/2025.02.04.25321392

**Authors:** Helen Anderson, Louise Brady, Joy Adamson

**Affiliations:** York Trials Unit, Department of Health Sciences University of York, York, United Kingdom YO10 5DD; General Practice Nurse (Locum) North Yorkshire; National Primary Care Nursing Lead (England) Nursing Directorate, NHS England 5th Floor (5W33), Quarry House Quarry Hill, Leeds, LS2 7UE

**Keywords:** General Practice Nursing, primary care, qualitative research, retention

## Abstract

**Background:** Increasing shortfalls in nursing workforces are detrimental to safety critical patient care. In general practice in England up to one-in-two nursing posts are predicted to be unfilled by 2030/31, with Wales similarly threatened. This is reflected internationally. Limited attention has been paid to how cultural and structural issues affect retention of nurses in general practice. The aim of our study is to understand factors that challenge retention and support nurses to stay in general practice.

**Methods:** We conducted an exploratory qualitative interview study with n=41 with members of nursing teams working in, or who have worked in, general practice as well as nurse leaders associated with general practice across England and Wales. Recruitment was through professional and social media networks and snowballing techniques. Data were collected between October 2023-June 2024 and analysed following framework analysis. University of York ethics approval (Ref: HSRGC/2023/586/A) was gained. The study was funded by the General Nursing Council Trust.

**Results:** Recognition of value of nurses working in general practice was central to the retention of nurses at all levels of practice and was affected by structural and cultural issues and reflected in several themes: The essence of nursing in general practice; The commodification and deprofessionalisation of nursing in general practice; Opportunities for development; Employment of nurses outside of the National Health Service; Lack of voice, precarity of position and lack of recourse; Tipping points.

**Conclusion:** Cultural and structural issues impacted on retention of nurses in general practice. While some supported retention, others revealed deep-seated, complex issues which require addressing at practice, local and national organisational levels. Nurses in general practice experience factors which leave them vulnerable and underserved. Policy makers, employers and professional organisations need to work to support retention and enable nurses in general practice, not only to survive, but thrive.

**Protocol Registration:** Open Science Framework (https://osf.io/) Identifier: DOI 10.17605/OSF.IO/2BYXC https://osf.io/2byxc/

## 1. Background

Increasing shortfalls in nursing workforces internationally are detrimental to the provision of safety critical patient care^1^. In England, one-in-ten nursing posts are left unfilled ^2^, while in general practice between one-in four and one-in-two nursing posts are predicted to be unfilled by 2030/31^3^ with 28% of nurses considering leaving general practice within the next year^4^. Nurses in general practice in England deliver 84 million patient contacts per year and provide cost-effective improved outcomes for patients, practices, wider communities and the National Health Service [NHS] more broadly ^5^. In Wales, the future of primary care is considered ‘at risk’ due to an ageing general practice nursing workforce^6^, with 20% being ≥60 years old^7^. These shortages of primary care nurses are reflected internationally^8,9^.

Issues identified as negatively impacting on the general practice nursing workforce include: inequitable pay, terms and conditions; lack of visibility, value and recognition of the complex nature of nursing in general practice nursing^10,11,12,13,14^; lack of nursing involvement in higher level decision-making^10,15^ and entrenched clinical and managerial hierarchies and hegemony^10^. Intention to leave general practice is associated with lack of respect, support and poor employment terms and conditions^10,16^.

The shape of general practice is changing with role diversification becoming increasingly prominent within primary care^2^. In England in 2019, NHS England (which is responsible for workforce planning) introduced the Additional Roles Reimbursement Scheme (ARRS) to general practice^17^. Healthcare practitioners who are not General Practitioners [GPs] or Registered Nurses [RNs], such as pharmacists or paramedics and physician associates, as well as nursing associates, are employed to take on work traditionally within the remit of registered nurses. These roles are centrally funded, and are employed at a Primary Care Network level, so have less financial cost to general practices^17^. However, the extent to which these issues affect retention of nurses working in general practice is not well understood and limited attention has been paid to retention despite a rise in healthcare professionals leaving the healthcare workforce more broadly^18^. Little research attention has been paid to factors which may support or challenge retention of nurses working in primary care internationally^19^. It is therefore important to consider the impact of cultural and structural issues on retention of nurses working in general practice.

In this paper we explore the cultural and structural issues associated with attrition and retention among nurses in general practice. Factors which may mitigate or exacerbate retention issues and identify factors to support retention are identified with a view to supporting employers and policy makers in future primary care workforce planning.

## 2 Methods

### 2.1 Aim

To understand underpinning cultural and structural factors that challenge retention and identify support for nurses to stay in general practice.

### 2.2 Objectives

To explore factors associated with retention of nurses in general practice, intention to quit and changes in professional plans.

To identify and explore how, and in what ways, general practice culture and structure influences, and is associated with, retention

To explore factors which may support or challenge general practice nursing workforce retention to inform future policy and practice.

To develop key factors to address attrition to support employers and policy makers in future primary care workforce planning.

### 2.3 Study Design and Setting

An exploratory qualitative interview study^20^ was conducted among nurses working in general practice across England and Wales and national leaders, with data collected between October 2023-June 2024. The study was underpinned by a social constructionist perspective to gain in-depth understanding of the effects of workplace culture and structure on retention of nurses working in general practice. It was funded by the General Nursing Council Trust [approved July 2023] for a 12-month period from 1 September 2023.

### 2.4 Participants

Nurses working in, or who had recently left, general practice in England and Wales at different professional levels (general practice nurses [GPN], health care assistants [HCA], advanced nurse practitioners [ANP], trainee Advanced Nurse Practitioners [tANP], nursing associates [NA], assistant practitioners [AP], nurses in management/leadership positions) and national leaders were eligible to take part.

### 2.5 Recruitment

Participants were recruited via professional and social media networks^10,21^. The lead researcher [HA] posted an explanation of the study on ‘X’ [formerly Twitter] and key groups with a strong and diverse nursing presence, such as @WeGPNs, @RCNGPNForum, @BAMEGPNs and @TheQNI, were asked to share information with the intention of gaining maximum variation of representation. Variation was also obtained by snowballing^22^ professional networks of clinical contacts nationally and via communications which targeted specific groups (e.g. to ensure black and minority ethnic representation, nurse leaders highlighted the study in particular geographical areas). A professional journal for nurses working in general practice also published details about the study which led to additional recruitment.

### 2.5 Sampling Strategy

The sampling strategy aimed to achieve a varied sample, balancing breadth and depth of data^23,24^. Recruitment was phased. We recruited opportunistically initially, with participants asked to complete a questionnaire of key personal and workplace characteristics. We then targeted specific groups based on gaps in recruitment (e.g. we snowball sampled health care assistants as this group did not respond to initial promotion of the study). It was anticipated that approximately 30–40 interviews would provide appropriate depth and breadth of data^25^. The demographic data were particularly important in selecting participants for maximum variation. Potential participants were given an information sheet and written informed consent was obtained from all taking part. Participants were thanked with a gift voucher.

### 2.6 Data Generation

Semi-structured interviews were conducted by the lead researcher [HA] at participants’ convenience via Zoom/MS Teams or via the telephone and lasted between 45–90 minutes. These were guided by an iterative topic guide underpinned by current literature and study aims and objectives^24^. Interviews were audio-recorded and transcribed verbatim by a professional transcription service and then audio files deleted.

### 2.7 Data Analysis

Data were analysed by the lead researcher [HA] using a framework approach^24^ consisting of: familiarization with the data; constructing a thematic framework; labelling and sorting (coding) the data; reviewing data extracts; data summary and display; abstraction and interpretation. This involves systematic analysis of qualitative interview data drawing on concepts developed from the research question/objectives as well as being grounded in the raw data. A constant comparative approach was taken. Data were collected and analysed concurrently enabling prospective themes and relationships to be tested in the data. The data set was searched for negative cases and alternative explanations.

### 2.8 Ethical Considerations

Research governance approval was gained from University of York Research Governance Committee (Ref: HSRGC/2023/586/A) on 29th September 2023. Data were held in accordance with the General Data Protection Regulations, the Data Protection Act (2018)^26^ and the University of York’s Data Management Policy. Unique identifying numbers pseudonymized identifiable data. Due to the relatively small number of participants and detailed data collection, the information presented limits potential for identification.

### 2.9 Rigor and reflexivity

Reporting is per COREQ^27^. We sought ‘naturalistic generalizability’, where findings are recognizable to those with shared experience^28^. Holloway’s understanding of rigour in qualitative research was followed by developing thick description, linking findings to theory, comparison to prior work and resonance with experiential knowledge^29^.

Both the lead researcher [HA] and another team member [LB] are registered nurses with experience in general practice and this had the potential to impact on the research process. Consequently, a reflexive approach was taken, and regular meetings held within the research team, which was also made up of a non-clinical methodologist [JA]. This supported sharing and questioning of analytical ideas, while alternative explanations were pursued.

## 3. Findings

### 3.1 Characteristics of participants

41 participants were interviewed across England and Wales (England n=35; Wales n=6. Details of participants are set out in Table 1. Most were female (n=35) with n=6 male. Most participants described themselves as white (n=32) and aged between 40-60 years (n=24). Participants worked across a variety of practices in terms of practice size, region, rurality and practice population deprivation scale (Table 2). We interviewed a range of nursing team members including GPNs, ANPs, HCAs, NAs and APs, as well as local and national nurse leaders from both England and Wales. A group interview also took place with two non-nurses, recruited through snowball sampling, who were working to support retention of nurses in general practice in a specific area of England. Participants ranged from those new to nursing to those who had retired. They held a range of qualifications up to and including master’s level, including prescribing qualifications. N=18 had been registered nurses for over 20 years. Participants varied from those who were currently working in general practice, to those considering leaving or actively looking for other positions and those who had already left general practice or retired. N=3 had retired early and returned to different primary care roles.

**Table 1:** Participant Characteristics.

| **Participant Demographic Data n (%)** |  |
| --- | --- |
| **Gender** |  |
| **Female** | 35 (85.4) |
| **Male** | 6 (14.6) |
| **Ethnicity (self-defined)** |  |
| **White British** | 25 (61) |
| **White Irish** | 2 (4.9) |
| **White Northern Irish** | 1 (2.4) |
| **White European** | 1 (2.4) |
| **White Welsh** | 3 (7.3) |
| **Black British** | 1 (2.4) |
| **White Indo-Caribbean** | 1 (2.4) |
| **Black** | 1 (2.4) |
| **British Indian** | 2 (4.9) |
| **British Asian** | 1 (2.4) |
| **Mixed** | 1 (2.4) |
| **Black African** | 1 (2.4) |
| **Mixed White** | 1 (2.4) |
| **Country of Work** |  |
| **England** | 35 (85.4) |
| **Wales** | 6 (14.6) |
| **Age range** |  |
| **20-29** | 4 (9.8) |
| **30-39** | 8 (19.5) |
| **40-49** | 12 (29.3) |
| **50-59** | 12 (29.3) |
| **60-70** | 5 (12.2) |
| **Qualifications** |  |
| **RN (RGN)** | 7 (17.1) |
| **RN (Dip)** | 5 (12.2) |
| **RN (BSc)** | 16 (39) |
| **RN (RSCN)** | 1 (2.4) |
| **RN (MSc/PGDip)** | 2 (4.9) |
| **RN (NOT STATED)** | 4 (9.8) |
| **ANP MSc/ PGDip** | 9 (22) |
| **NMP** | 10 (24.4) |
| **Foundation degree (NA/AP)** | 4 (9.8) |
| **HCA Level 3/4** | 3 (7.3) |
| **Other MSc/MA** | 7 (17.1) |
| **Other Batchelor's degree** | 4 (9.8) |
| **Other post grad quals** | 9 (22) |
| **N/A (not a nurse)** | **2 (4.9)** |
| **Years Qualified** |  |
| **1-9** | **6 (14.6)** |
| **10-19** | **6 (14.6)** |
| **20-29** | **5 (12.2)** |
| **30-39** | **9 (21.2)** |
| **40-49** | **4 (9.8)** |
| **Not stated** | **2 (4.9)** |
| **RNA/AP** | **1-9YRS: 3 (7.3) 10-20 YRS: 1 (2.4)** |
| **HCA** | **1-9 years: 3 (7.3)** |
| **N/A (not a nurse)** | **2 (4.9)** |
| **Role** |  |
| **GPN** | **10 (24.4)** |
| **ANP** | **6 (14.6)** |
| **RNA/AP** | **4 (9.8)** |
| **HCA** | **3 (7.3)** |
| **Trainee ACP (currently GPN)** | **4 (9.8)** |
| **GPN/ANP combined role** | **3 (7.3)** |
| **Nurse Leader** | **7 (17.1)** |
| **Other PC role** | **1 (2.4)** |
| **N/A** | **2 (4.9)** |
| **Employment Status** |  |
| **In general practice** | **23 (56.9)** |
| **Left GP** | **3 (7.3)** |
| **Retired (early)** | **1 (2.4)** |
| **Retired early and returned (different role)** | **3 (7.3)** |
| **Left for another role in PC** | **2 (4.9)** |
| **Nurse Leader** | **7 (17.1)** |
| **Not applicable** | **2 (4.9)** |
| **Pay/Agenda for Change [NHS] equivalent (2024)** |  |
| **Band 3 (£11.67-£12.45/hour)** | **2 (4.9)** |
| **Band 4 (£12.86 -£14.21)** | **6 (14.6)** |
| **Band 5 (£14.53-£17.69)** | **2 (4.9)** |
| **Band 6 (£18.10-21.80)** | **6 (14.6)** |
| **Band 7 (£22.37-£25.60)** | **9 (22)** |
| **Band 8a (£26.06-£29.33)** | **5 (12.2)** |
| **Band 8b (£30.16-£35.04)** | **1 (2.4)** |
| **Band 8c (£36.01-41.50)** | **1 (2.4)** |
| **Band 8d (£42.74-£49.29)** | **1 (2.4)** |
| **Band 9** | **2 (4.9) (Nurse leaders)** |
| **N/A** | **5 (12.2)** |
| **Not stated** | **1 (2.4)** |

**Table 2:** Practice Descriptors.

| **Practice Descriptors (32 practices)** |  |
| --- | --- |
| **Small (<10k)** | 14 (43.8) |
| **Medium (10-20k)** | 15 (46.9) |
| **Large (>21k)** | 2 (6.3) |
| **No Info** | 1 (3.1) |
| **N/A** | 9 |
| **Rurality** |  |
| **Rural/semi-rural** | 6 (18.8) |
| **Suburban** | 5 (15.6) |
| **Urban** | 16 (50) |
| **Mixed** | 3 (9.4) |
| **Other** | 1 (3.1) |
| **No info** | 1 (3.1) |
| **N/A** | 9 |
| **Region** |  |
| **England (NE)** | 0 (0) |
| **England (NW)** | 6 (18.8) |
| **England (Yorkshire)** | 4 (12.5) |
| **England (Midlands)** | 4 (12.5) |
| **England (SE) inc London** | 12 (37.5) |
| **England (SW)** | 0 (0) |
| **Wales** | 5 (15.6) |
| **No info** | 1 (3.1) |
| **N/A** | 9 |
| **Practice Population Deprivation Level\*** |  |
| **1 (most deprived)** | 2 (6.3) |
| **2** | 5 (15.6) |
| **3** | 6 (18.8) |
| **4** | 3 (9.4) |
| **5** | 0 (0) |
| **6** | 3 (9.4) |
| **7** | 0 (0) |
| **8** | 4 (12.5) |
| **9** | 4 (12.5) |
| **10 (least deprived)** | 1 (3.1) |
| **No data** | 3 (9.4) |
| **N/A** | 10 |
| **All Practices (where applicable) were CQC rated Good except 1 which was rated requires improvement and 1 rated outstanding** |  |

\*National General Practice Profile data (https://fingertips.phe.org.uk/profile/general-practice) or https://statswales.gov.wales/Catalogue/Health-and-Social-Care/General-Medical-Services/General-practice-population

### 3.2 Overview

Recognition of the worth and value of nurses working in general practice is central to the retention of nurses at all levels of practice and is affected by structural and cultural issues. Several themes were developed: The essence of nursing in general practice – variety, continuity and holism; The commodification and deprofessionalisation of nursing in general practice; Opportunities for training and development; General practice employment of nurses outside of the NHS; lack of voice, precarity of position and lack of recourse, and tipping points. Each theme, and associated sub-themes, will be explored in turn.

### 3.3 The essence of nursing in general practice – variety, continuity and holism

There were several positives which drew and kept participants working in general practice. They valued the diversity and variety of general practice work, providing care across the lifespan and for patients with a vast range of physical and mental health conditions. They are at the forefront of health promotion and public health, delivering complex childhood and other immunisation and vaccination programmes, screening programmes, provision of women’s health, consulting on travel health and providing expert wound care. Many also managed long term conditions.

> I do enjoy patient facing [work]. I love hearing about people’s stories, different backgrounds. You might have somebody that [needs] extensive travel advice and in the next consultation you will have a baby immunisation and the next one you might have somebody for an ECG, so I like how varied it is across all ages. [ID 128 ANP working as GPN, female]

Participants enjoyed the challenge of their complex workload, and this was considered central to retention, even if this level of practice was not always recognised, *‘I hate the word general practice nursing because that’s quite derogatory. Practice nursing is specialist nursing. You have to know all the areas [ID 102, GPN/tANP male].* Nurses at all levels valued continuity of care, being able to develop trusted and therapeutic relationships with patients, and often families, across generations. They also understood the local context of practice populations. This all led to high levels of job satisfaction and enjoyment of their work, *‘you could build those relationships with your patients…. follow their journey and you’re part of it’ [ID107 GPN, male]*.

### 3.4 Commodification and deprofessionalisation of expert nursing

However, there was a consensus that these positive aspects could be lost by salami slicing and reduction of care to tick box exercises and pressure from employers to do more with less. This contributed to what some saw as the commodification of nursing in general practice and the shift towards a deprofessionalisation agenda, which was considered to be negatively implicated in retention.

> it’s the pressure from the GP partners to get more people in, to tick more boxes. Our appointment times were reduced with no consultation whatsoever….So, a lot of the consultations, the clinical focus can sometimes be overridden.

> (ID 106 GPN, Female)

This was noted by many to be exacerbated by the introduction of new roles into the primary care nursing space. HCAs and APs are established members of the general practice nursing team. In England (but not currently in Wales), the Nursing Associate role was introduced to bridge the gap between Registered Nurses and Health Care Assistants (Willis, 2015)^30^. While those working in these roles were valued members of the nursing team, at some practices, NAs and HCAs were taking on work previously within the domain and remit of registered nurses and this has implications for retention.

> I run my own clinics and do everything that a nurse would do apart from travel vaccinations and child imms. Advanced wound care, smears, injections, falls, everything.…I do diabetic reviews. I do newly diagnosed diabetic patients. I do asthma reviews.

> [ID 103 NA, female]

This was seen by some to reduce nursing care to a series of tasks, hence undervaluing the role and level of practice, posing a risk to patient safety and decreasing job satisfaction. It was considered that knowing how to carry out a task was being conflated with having the professional knowledge and acumen to use that information to inform clinical judgment,

> *‘no disrespect to nursing associates, they don’t have the knowledge and skill at that registered nurse level…. people don’t realise or will realise it too late [ID 102 GPN/tANP male]*.

The Nursing and Midwifery Council [NMC], which regulates and sets proficiency standards for both registered nurses and registered nursing associates in England^31,32^, states that NAs’ scope of practice is defined at a local level by the employer^33^, who in most cases in general practice are GPs or non-clinical managers. This meant that while registered nurses were professionally responsible for NAs’ work, they often had little access to the decision-making underpinning the delegation of that work. It also meant that HCAs and NAs were potentially at risk of exploitation, may ‘not know what they don’t know’ and could be left without adequate RN supervision.

> this is about bolstering a nursing workforce through a side door, instead of investing money in nurses. Nursing associates are being used unsafely, working in practice without a registered nurse being employed, which goes completely against the governance behind the NA role. I think it’s being exploited.

> There is a huge danger that the value of the RN role is being completely missed

> [ID 126 GPN and nurse partner, female]

However, many registered nurses actively supported the developing scope of support workers, arguing that there was more than enough work to go around. It was considered by some that GPNs needed to ‘up their game’ to further develop their level of practice. It was highlighted that registered nurses did not always see the bigger picture or recognise their role and responsibilities in leading and influencing the agenda around nursing associates.

> If we link it back to retention, nurses start to feel undervalued because they think [employers] can just get an NA to do [the work]. They don’t take authority in their role. They give away their power. There is something in the mind-set of the general practice nurse themselves that needs to change in taking on that leadership role and articulating that.

> [ID 203 National Nurse Leader, male]

Participants also felt practice owners used Additional Roles Reimbursement Scheme (ARRS) roles, rather than employing registered nurses, as it was less costly to practices. This was considered another factor negatively affecting nurse retention as it highlighted a perceived lack of recognition and value of experienced and knowledgeable registered nurses working in general practice.

> we’ve got groups of PAs [Physician Associates] that are now doing long-term condition[s] and nurses are back to what I describe as more generalist treatment room nurses. [ARRS has] definitely had an impact, particularly on the larger practices in our PCN. A huge motivating factor is the benefit financially that has brought to practices, then they just change everything else around that. Where they’ve lost their more experienced practice nurses, they’ve chosen not to replace them and then when practice nurses have come in, they’ve felt as though they’ve been unsupported. They don’t feel valued as health professionals.

> (ID 101 Ex GPN now PCN lead, female)

Differences in remuneration between people employed on ARRS contracts, and nurses employed directly by general practice, highlighted further perceived inequity and lack of value placed on nursing in general practice, *‘[a paramedic] told me how much he was going to make and I think that was maybe [£s] more than what the nurses were on….you do feel a bit like “you’re not valuing me, what I’m doing here?’ [ID 131 GPN, female].* Indeed, the development of ARRS, and the way it was introduced with little or no consultation with nurses working in general practice, and how this might affect their role, was seen to create a divisive atmosphere which further reduced nurses’ feeling of being valued. Both the introduction of ARRS and the development of NAs and HCAs led to some registered nurse participants feeling pushed out of general practice, with participants unsure whether this is an unintended consequence of poor policy, or a deliberate attempt to undermine nursing, *‘Is that what the future holds - the snipping away at the practice nurse role? [ID 120 GPN retired and returned, female]*.

### 3.5 Opportunities for training and development

A positive aspect of working in general practice which was considered to support retention was the potential for career progression and development, and educational opportunities, in comparison to working in secondary care.

#### 3.5.1 ‘Grow your own’

Participants reported being encouraged and supported to undertake specialist and postgraduate qualifications, *‘[The training] in primary care is abundant and [you] learn so much…The educational side is excellent….I think these are things that can help retain staff. [ID 115 GPN, Female].* As well as supporting registered nurses to develop, practices were often supportive of ‘grow your own’ development which meant that members of the reception team, HCAs, APs and NAs were supported to develop their roles, *‘I started out working in reception and then they trained me to a HCA role….then there was an opportunity [to do] the assistant practitioner [training]’. [ID 125 Assistant Practitioner, female]*

One potential pathway for doing so was via the introduction of apprenticeship routes. Apprentices are employees who undertake funded education alongside their work and apprenticeship education programmes are available at nursing associate, registered nurse and advanced practice levels. These development routes were singled out as a positive way to support people to develop their career within general practice and therefore retaining staff who want to progress. This was seen as a way of retaining nurses within general practice longer term.

> Topping up from nursing associate to registered nurse through the apprenticeship route should hopefully be completed by May and I’m just going to carry on at [practice name] for a while and maybe go on to do the prescribing course and maybe, if I get the chance, to do the ANP apprenticeship ….at [practice name] my confidence has been really built up and I’ve been encouraged to do all these things.

> [ID 124 NA, male]

Apprenticeships were considered to widen participation and support people who, for various reasons, may not have been able to undertake traditional nurse education. It was also considered to support retention because it targets people who live locally and are more likely to have commitments which would keep them in local employment. However, development opportunities were variable and dependent on individual employers’ support. For example, some participants were expected to find their own external training placements required for apprenticeships or were not allowed time off. Upskilling of staff was also not necessarily adequately recognised and rewarded by practices, leading to participants feeling discontent and sometimes taking their labour elsewhere. Some participants were clearly ambitious in terms of career development and saw general practice as a stepping stone, where they could gain expert skills and knowledge, and then leverage this to gain roles with greater remuneration both inside and outside general practice.

#### 3.5.2 The (re)conceptualisation of advanced practice

Registered nurse participants felt that the only way they could progress in terms of career, status and, especially, remuneration in general practice was to become an ANP. While this was seen as an important development opportunity by some, it was also considered to negatively affect retention. NHS England funding of advanced clinical practice apprenticeships, where nurses were educated to carry out work more traditionally undertaken by GPs, was seen to be prioritised over other nurse education. The primacy of this kind of advanced practice resulted in some participants feeling that nursing in general practice more broadly was being devalued. It was also dissonant with some participants’ philosophy of nursing, with some feeling they were losing the essence of nursing, *‘I was no longer a mega nurse. I was expected to replace GPs basically and that was never my intention’ [ID 137: ANP/GPN Fellowship Lead Nurse, Female].* Because advanced practice was seen in terms of managing patients with undifferentiated diagnoses, and closer to the medical model of GP work, this was valued more highly than general practice nursing work. Participants wanted to develop, but not necessarily as ANPs. However, the structural framework was not in place to support this. GPNs saw the breadth and depth of their work as that of a specialist generalist with equivalence to advanced practice, but felt GP employers and managers did not recognise the level of practice at which GPNs worked. Despite publication of documents mapping nurses’ level of practice from support worker to consultant level^34^, such interventions were not considered effective as employers were not required to implement them.

*As a practice nurse, I pushed that [GPN] role as far as I could*.

> I did the RCN [Royal Collage of Nursing] Accreditation. I became a non-medical prescriber. I did degrees in every chronic disease. I didn’t want to be an ANP because I see the roles as totally different…There was this new structure that came out with general practice nurse consultants but it’s not being adopted widely….I’m not sure if practices want it because I think they’re happy for practice nurses to be working at an advanced level without the recognition and without the increasing pay.

> [ID 120, GPN, female]

### 3.6 General practice employment of nurses outside of the NHS

Reflecting the national picture in England and Wales, nurses working in general practice in this study were employed, in the main, by individual general practices. These are usually owned and managed by general medical practitioners, and sometimes practice managers, who form financial partnerships. These partnerships provide services for, but sit outside, the NHS. This means they set out their own individual employment conditions. Whilst they are required to follow agreed remuneration packages for salaried GPs, they set their own salary and terms and conditions packages, such as sick and maternity pay and annual leave, for other employees. Being employed outside the NHS meant that the experiences of nurses working in general practice were different to other nurses and this had its own benefits and challenges which were often in tension with each other.

#### 3.6.1 Flexibility trade off

An aspect of working in general practice considered to support nurse retention was the potential for flexibility and family/life-friendly hours. Many participants reported being initially attracted to general practice as it allowed them to develop their professional career alongside caring and other responsibilities, within a close geographical area, without the need to work unsocial hours. Because participants were employed by small organisations, with access to decision-makers, many nurses in our study found that there was more flexibility in negotiating workarounds for things like attending school events or caring responsibilities. However, this informal flexibility and close geographical proximity meant that nurses often had minimal scope to take their labour elsewhere. They felt that poor pay and/or terms and conditions were a trade-off for being able to manage daily life and work. However, this tended to be time-limited and served to hinder retention longer term.

> Surgeries can pay whatever they want and, when I first went there, I took a pay cut, but you accept it at that point [because] I’d not done practice nursing before and it’s a trade-off. I don’t have to work weekends or bank holidays. I get Christmas off. So that was quite attractive. But then as you get further on, and you do things like the prescribing [course], and don’t even get offered a pay rise. You’re doing this massive important role, and you get very little recognition

> [ID 104 Ex GPN, Female]

#### 3.6.2 Association between feeling valued and remuneration, terms and conditions

Participants highlighted that the contribution they made, and their level of practice, often went unrecognised by their GP employers and practice managers. One of the main ways this was illustrated was through remuneration and associated terms and conditions, which were seen as markers for professional respect, *‘I think a huge part is recognition and the other part is financial’ [ID 102 GPN/tANP male]*.

Employment within individual general practices meant participants were required to negotiate their own employment packages and this could be problematic in several ways. First, it required nurses to negotiate directly with their employers, with whom they often have a close working relationship. This could cause tensions when they felt their worth was not valued and sour collegiate and employee-employer relations. In other healthcare employment structures in England and Wales, such as NHS Trusts, nurses are employed on nationally standardised Agenda for Change terms and conditions. Consequently, many participants had little experience or knowledge about how to negotiate their worth and struggled to both articulate this and negotiate effectively, *‘you have to negotiate your salary and I was very much uncomfortable with that. I didn’t know - am I asking for the right amount?’ [ID 134, GPN and Federation Lead, Female].* Secondly, nurses were often met with challenge, or were surprised when they were not, based on past experiences, *‘It’s a shame that you have to battle and fight for everything that you want and practice nurses deserve. When you look at the responsibility and what’s involved in the job [ID 107 GPN, male].* Participants were often required to justify pay rises and were sometimes actively moved against when they asked for recognition of their worth. One participant who worked as a GPN, but had now left this role, illustrates the complexity and difficulties nurses experience and how remuneration is intrinsically linked to feeling valued. For this participant, lack of recognition resulted in them leaving general practice nursing.

> One of the final straws was my [last] appraisal. I never ask for a pay rise.…I laid all the skills out that I’d gathered over the years. I’d never had a negative appraisal. I said ‘I know [in] general practice there isn’t a big pool of money. However, I feel like I need something to at least show that you’re recognising what I’ve done’. And it basically got kicked back and, ‘well you need to show us that you’re safe and competent. We want someone to sit in with you’. I’d already been performing these reviews and at no point had anyone, no patient complaints, no GP, had said to me, ‘you’re not doing this right’ I couldn’t keep coming home loving what I did for the patients but feeling not really sure why I was doing it for that practice….It felt very much like attitudes almost changed a little bit towards me.

> [ID 117 Ex GPN now moved to another role, female]

At the time of data collection, practices had been advised by the government to uplift practice staff’s pay by 6% and were provided, to some extent, with funding associated with this. However, practices were not compelled to do this and the majority of nurses in the study reported that they did not get any or the full uplift, or it had not been discussed with them, or the decision had been deferred to a later date.

Consequently, participants felt employers actively did not want to pay them what they were worth, and this directly influenced their intention to quit.

> Even with the 6% I’d still get underpaid. I don’t feel valued at all and the only reason I’m still there is to finish my [ANP] training, then I will move on….I will not be going anywhere close to primary care unless there is pay and conditions. [ID 123 GPN/tANP, female]

Participants considered that employers almost engaged in a wilful blindness to the work of nurses in general practice because recognising their worth would mean that remuneration would need to adequately reflect this and this, in turn, would affect their own finances, ‘*If they can get away with it, and they can put an extra fiver in their pocket, they’ll do it’ [ID 102 GPN/tANP male].* Related to this, annual leave entitlement and sick and maternity pay, was also decided at a practice level, with many participants receiving below Agenda for Change conditions, *‘maternity leave, which is a big thing for me, is just statutory and there is nothing you can do to change that even when you are trying to have negotiations and the same for sick leave’ [ID 128 ANP, Female].* As well as the obvious issue of this contributing to nurses choosing not to join or stay in general practice; that many practices chose to pay highly skilled professionals only statutory sick and maternity pay again contributed to nurses feeling devalued as a professional group.

> there’s such a disconnect between how GPs see their own income relating to pay, terms and conditions and everybody else can go to hell, frankly, on a micro level. GPs as a profession have an absolute blind spot over how they treat employees….Which other professions pay statutory sick pay for very expensive well-trained highly qualified people?

> [ID 105 Nurse Leader, Female]

For their part, before joining general practice, many participants assumed they would be employed under Agenda for Change terms and conditions. Some participants remained unaware of their entitlements even after taking up employment, *‘that’s not something I’m 100% sure on because I’ve never needed to be off sick. And then with maternity I’m again not 100% sure how that would work’ [ID 135 HCA, female].* For participants with long term or complex medical issues, this could be particularly problematic. It was felt that employers could act with impunity with little recourse for their actions and the nature of general practice meant that nurses experiencing difficulties with their employers felt isolated and unsupported.

> HR policies vary so much and there is no [set process like in] an NHS Trust. I had a very bad year of severe sickness and there wasn’t any support. I’ve been asked to attend meetings and then I’d be confronted with HR managers [and] partners.

> …that was just not a way to deal with a colleague. I think the trust goes [and] people are quick to leave

> [ID 128 ANP/GPN, female]

Furthermore, it was thought that isolation was a tool used by employers to prevent nurses uniting or using their voice collectively. Some participants suggested that GP employers deliberately tried to keep nurses isolated from nurses at other practices, local Primary Care Networks [PCNs] and Integrated Care boards [ICBs] to prevent them knowing other nurses’ terms and conditions or acting collectively, *‘[practices] don’t want them to mingle with other nurses because they might share ideas, they might [discuss], “how much are they paying you?”[ID 134 GPN and Federation Nurse Lead, female]*.

### 3.7 Lack of voice, precarity of position and lack of recourse

Participants felt isolated from their professional associations. While they felt supported at an individual level (e.g. if they were experiencing bullying), they did not feel adequately represented as a professional group. Despite, for example, recent work by the RCN supporting general practice nursing terms and conditions^35^, this was not felt ‘on the ground’. Participants thought professional organisations were unable to effectively negotiate for nurses working in general practice, partially due to their unique employment status. This was in contrast to the perception of support for GPs from medical organisations. While participants themselves often did not engage in the political or professional arena, professional organisations were perceived to be ineffectual, leading to a weakened professional status and lack of a strategic level voice for nurses working in general practice.

*They’re not particularly useful in advocating for the profession*.

> I don’t think they’re [the RCN] very successful in their outcomes, so we’ve not really got a voice at Government level. The GP businesses will make their own decision[s]. At a local level nurses don’t particularly work well together, they’re not very political. You can then look at the union representation at Government level and that is pretty rubbish, so in general our voice is quite weak.

> [ID 122 ANP, female]

Participants were, in the main, frustrated that they were unable to contribute to decisions about their work at practice and wider organisational levels. They felt lip service was paid to nurses’ views, leaving them feeling undervalued and negatively impacting retention, *‘They like to make me think they’ll take it on-board. But then the practice manager has a tendency to [say], “well no, we all think this is what you should be doing” and that’s that.’ [ID 104 Ex GPN, female].* Participants recognised that the situation with their employers could suddenly change, sometimes with a change of partner or manager, but often without identifiable reason - highlighting the precarity of their position. This led to nurses moderating their behaviour. They chose their battles and, because they saw patient care as a priority, would speak up for that at the expense of their own wellbeing. This has implications for retention.

> You can voice your opinion but it’s in a guarded way because they are your employer. In a hospital setting, you would be able to say what you wanted because there’s no consequence in terms of your employment. But if you’re being employed by these people, then they have a lot more control over you and you don’t want to damage your employment history with them by voicing things that they may not like….There’s a massive part of organisational culture involving retention.

> [ID 122 ANP, female]

That it was necessary to be cautious was borne out by some participants who had experienced bullying when challenging GP employers. Consequently, nurses had to know how to ‘play the game’ to get their voices heard without upsetting the status quo. There was recognition that the model of general practice was weighted against nurses having a voice and sometimes the only option was to move on.

> I am looking for another practice. There is an element of bullying from one of the partners. Because I stood my ground, she really didn’t like it. …. At the same time, I don’t want to jump ship and find myself somewhere even worse!

> [ID 106 GPN, Female]

A minority of participants did feel they had a voice at a strategic and decision-making level. This was seen as important because, not only were nurses’ views represented, there was recognition that general practice nursing as a profession was valued. This is exemplified by one GPN who was a practice partner. However, this was very much a minority position.

> the existing partnership recognised what I was bringing to the team, and how that could complement what the partnership offered. Nursing leadership actually is very different from medical leadership…and I think they recognise the value of having that in the partnership. They are a very progressive team versus some of my colleagues around the country. [ID 126 GPN and nurse partner, female]

### 3.8 Tipping points

Participants who had left general practice, or were actively seeking employment elsewhere, often spoke of a ‘final straw’ or tipping point which led them to this decision, *‘It was the checking up on things which was insulting. The straw that broke the camel’s back after the role that I’d been doing well’ [ID114 Retired GPN, female.]* If one participant left their practice, this often either prompted others to leave, or was part of a pattern of high turnover, which indicated nurses did not feel valued or well-treated, *‘I only stayed there for a year because I didn’t have a good time. When they hired me, they hired two other nurses, but they left [ID121, GPN/tANP, female].* While nurses with long careers and those new to nursing often felt differently in terms of loyalty to general practice(s), there were limits to their tolerance relating to whether their worth was recognised and reached tipping points where they moved to take back ownership of what they considered to be devaluing of nursing and sometimes exploitation, *‘your tolerance for being able to put up with things lessens the nearer you get to retirement [ID 101 Ex GPN, PCN Lead Nurse, female].* It is therefore necessary for practice leaders to proactively recognise and reward the value of nursing’s contribution to general practice and acknowledge the worth of individual nurses.

> My generation, we’re not going to stay loyal if we’re not getting that mutual relationship, and remuneration is part of that process [of] value. If you don’t feel valued, I’m not prepared to stay and negatively impact on myself, my well-being, my career progression.

> [ID 107, GPN, male]

Others mentioned a lack of ‘good will’ extended to them as nurses who had contributed many years to general practice – leading to them reaching a tipping point which impacted negatively on retention.

> The final straw was being expected to arrange my father’s funeral for a day off, so that the surgery didn’t lose out… I’d been there for so long and the relationship I supposedly had with the GPs….What annoyed me was all the stuff I did for them when I could have been picking up my kids from school…and instead I was studying to get another course because [the practice] needed that.

> [ID 137 Retired ANP/GPN, female]

## 4. Discussion

Our study sample broadly reflects of the national demographic make-up of the general practice nursing population in terms of gender, age and ethnicity [Supplementary Table 1] with males slightly oversampled for maximum variation.

Findings indicate significant cultural and structural issues interact to impact on retention of nurses working in general practice. These relate to recognition of the value of nursing in general practice at all levels and areas of practice and their limited access and input into higher level decision-making. Also highlighted are the importance of development opportunities, the distinctive nature of the relationship between nurse and employer and the precarity this can produce, as well as a perceived weak professional representation and voice.

Positive factors supporting retention included variety and continuity of care, complexity of work, opportunities to develop and therapeutic relationships, as well as flexibility and family-friendly hours^8^. A concept analysis of nurse retention more broadly suggests that motivation is crucial to attrition decisions, with self-determination, interest, excitement and curiosity being central motivators, while personal factors such as caring responsibilities also contribute^36^. Intention to stay in an organisation is associated with high job satisfaction and organisational commitment which is associated with organisational culture, such as flexible working and support to develop^1^. Clearly, then, employers ought to maximise these positive elements as protective factors in retaining nurses in general practice.

> ‘Grow you own’ strategies and apprenticeship routes to NA, GPN and ANP education supported retention, widened access and employed a geographically local workforce in our study. Attachment to ‘place’ is an important aspect of retention and supporting local populations into nursing provides a sustainable local workforce^36^, with nursing apprentices likely to stay with their employer post qualification^38^. However, reflecting our study, lack of clarity about the NA role in terms of scope, remit, accountability, boundaries and professional identity have been identified previously^17^ while lack of support, scope creep and limited social mobility are disincentives to apprenticeships routes^37^.

Reflecting our findings, a recent Queen’s Nursing Institute [QNI] survey found the introduction of the Additional Roles Reimbursement Scheme led to disinvestment and devaluation of nursing work, increased ‘taskification’ of nursing care, role creep and inequitable pay and conditions, all of which have a negative impact on morale^38^. Like us, they found little consultation with nurses about the introduction of ARRS despite this having a significant impact on their work. Furthermore, unlike registered nurses in primary care, substitute roles lack evaluation in terms of patient safety^2^. Consequently, at policy and individual practice, Primary Care Network [PCN] and Integrated Care Board [ICB] levels, it is necessary for the impact of ARRS on general practice nursing workforces to be considered, and for nurses to be included in that decision-making, to support retention.s

We have highlighted that participants did not have a voice at senior levels of decision making, reflecting their lack of worth and impacting retention. Likewise, the Sonnet Report into the role and value of nurses in general practice in England^5^, associated attrition with feeling undervalued, being prevented from reaching potential and lack of contribution at leadership and decision-making levels. Decision-making contribution has been implicated in nurse vacancy rates more widely^36,39^.

Reflecting participants in our study, there are increasing calls for pay, terms and conditions of nurses in general practice to be commensurate with role complexity and consistent between practices^5,40^ though it is unclear how this will be achieved while business owners at a practice level continue to negotiate with individual employees. Our findings indicate some participants suspected their employers of ‘wilful blindness’ in their lack of recognition of the level of practice and complexity of work of nurses, in order to avoid commensurate remuneration and professional terms and conditions. This involves deliberate avoidance of facts or ‘turning a blind eye’^41^. However, terms and conditions alone are unlikely to significantly impact retention because it is linked to multiple factors including intrinsic motivation and ‘doing a good job’. This is challenged when the value of a professional group is disregarded, or when work is split into a series of tasks^18^.

As noted in our study, there is a significant association between bullying and turnover intention^42^. Amongst our participants, fear of bullying, or negative consequences of working in close proximity to their employer who did not appear to follow established employment practices, meant participants did not always speak up and their needs went unaddressed. Similarly, it has been found that social-relational aspects of medicine and nursing led to nurses feeling undervalued and professionally disrespected, resulting in nurses’ voices being silenced^43^. Lack of structural support in general practice is also an issue. For example, Freedom to Speak Up Guardians, who support whistle-blowers, were only present in slightly over 20% of primary care organisations in 2023/24^44^. Lack of a structured framework within which nurses’ rights are protected leave them in a precarious position.

Our findings indicate that even when relationships with employers were cordial, there was an underlying awareness that this was precarious, and participants needed to consider this in their everyday actions. When they stepped outside of demarcated boundaries by, for example, asking for a pay rise, they faced negative consequences. These issues negatively impacted on their enjoyment of work, contributed to silencing their voice and consequently affected retention. Furthermore, participants in our study felt disengaged with professional organisations, which they considered did not speak up for the specific nature of nursing in general practice. While professional organisations, such as the RCN, have more recently worked to support improved pay, terms and conditions for nurses working in general practice^35^, this did not appear to be the perceived experience of participants, who remained largely disappointed in, or unaware of, this work. Taken together, the employment of nurses in general practice is particularly precarious. Others have noted this is the case for the nursing profession more generally^45,46^ with nurses not benefitting from what is defined as ‘decent work’ including recognition, adequate income and conditions, security of position, contribution to workplace decision-making, opportunities to develop and a positive work atmosphere^45^.

Previous systematic reviews of nursing retention more broadly indicate pull-push factors likely to relate to age and career stage ^1,47^. Younger and less experienced nurses are likely to have lower attachment to their organisation and more likely to be transient^1^, while, reflecting our study findings, mid-career nurses related retention to lack of career advancement and late-career retention was associated with lack of recognition^47^. In our study there were some commonalities across age and experience groups, such as the requirement for workplace support, flexible working, opportunities for career advancement, fair treatment and remuneration and these were also identified across nursing generations more broadly^46^. However, our findings identified issues specific to nursing in general practice and indicated that participants reached a catalyst or tipping point of tolerance where previously accepted (or tolerated) ‘trade offs’ flip so that costs of working in general practice outweigh benefits. This led participants to seek to leave their current organisation, or general practice altogether. This usually revolved around lack of recognition, not feeling valued or listened to, and often followed a period of distress. Similarly, Holland et al^39^ suggest that lack of voice is a tipping point for exiting nursing. In our study, tipping points followed a recognisable pattern that could be potentially interceded to prevent attrition and support this group of professionals.

### 4.1 Strengths and Limitations

Sample homogeneity may occur in social media recruitment^48^. We countered this through a phased recruitment strategy, critically analysing ongoing recruitment using a demographic questionnaire, while also recruiting concurrently via professional networks. We were successful in recruiting a diverse range of nurses across England and Wales sampled for maximum variation in terms of gender, role, demographics of surgeries and geographical location, while age and ethnicity reflected the national picture.

A potential limitation was mainly interviewing nurses in general practice and nurse leaders. Data from a wider range of healthcare professionals, employers or managers/decision-makers and patients/carers might have provided alternative perspectives. However, funding and timeframe for the study were limited. Focusing on others may have detracted from depth and breadth of information from nurses themselves. The strength of qualitative research is in the information-rich data generated. This allows in-depth understanding of experiences of nurses working in general practice, the associations between retention and cultural and structural issues and how these can be addressed moving forward.

### Implications for policy/practice and recommendations for further research

Working in general practice is precarious for nurses and the distinct nature of their employment makes them a vulnerable, isolated and underserved group. As with any such vulnerable group, workforce policy, employers and professional associations have a responsibility to develop strategies to meet the needs of this highly skilled groups of professionals. Further research on the societal-level cultural and structural factors underpinning the issues raised in this paper is required to unpick how these are inculcated and impact on retention.

## 7. Conclusion

Cultural and structural issues were identified which impact on retention of nurses in general practice. While some supported retention, others challenged it and revealed deep seated and complex issues which require addressing at practice, local and national organisational levels. Nurses in general practice experience factors which leave them vulnerable and underserved. Policy makers, employers and professional organisations ought to work to support retention and enable nurses in general practice, not only survive, but thrive.

## Abbreviations

ANP: Advanced Nurse Practitioner [tANP= trainee]
AP: Assistant Practitioner
ARRS: Additional Roles Reimbursement Scheme
GPN: General Practice Nurse
GP: General Practitioner
HCA: Health Care Assistant
ICB: Integrated Care Board
NA: Nursing Associate
NHS: National Health Service
NMC: Nursing and Midwifery Council
PA: Physician Associate
PCN: Primary Care Network
RCN: Royal College of Nursing
RN: Registered Nurse
QNI: Queen’s Nursing Institute

## Declarations

### Ethics approval and consent to participate

Research governance approval was gained from University of York Research Governance Committee (Ref: HSRGC/2023/586/A) on 29th September 2023. Everyone who took part in the study had to fill out and sign an informed consent form at the start

### Availability of data and materials

The datasets generated and/or analysed during the current study are not publicly available due the being of an in-depth qualitative data set for a specific group of participants who could potentially be identifiable if whole transcripts were publicly available. Data sets are available from the corresponding author on reasonable request.

### Authors’ contributions

All authors contributed to the overall study and made substantial contributions to the conception and design of the work and in the acquisition, analysis and interpretation of data. HA drafted the paper. JA and LB regularly reviewed and commented the manuscript. All authors revised the text critically for intellectual content and approved the final manuscript.

## Data Availability

Data produced in the present study are available upon reasonable request to the authors

## Acknowledgements

The authors wish to thank participants for their valuable contributions to the study

## Authors’ information

[HA] and [LB] are registered nurses with experience working in general practice and this had the potential to impact on the research process. Consequently, a reflexive approach was taken. Regular meetings to share and question analytical ideas were held within the research team, which was also made up of a non-clinical methodologist [JA]. Throughout the process, assumptions were challenged and alternative explanations sought.

## References

1. Pressley C, Garside J. Safeguarding the Retention of nurses: a Systematic Review on Determinants of nurse’s Intentions to Stay. Nursing Open. 2023 Jan 16;10(5):2842–58. Available from: https://www.ncbi.nlm.nih.gov/pmc/articles/PMC10077373/

2. Dixon-Woods M, Summers C, Morgan M, Patel K. The future of the NHS depends on its workforce. BMJ. 2024 Mar 27;384(384):e079474. Available from: https://www.bmj.com/content/384/bmj-2024-079474

3. The Heath Foundation (2022). Projections: General practice workforce in England. www.health.org.uk. June 2022 https://www.health.org.uk/publications/reports/projections-general-practice-workforce-in-england

4. Ford M. Exclusive: 28% of GPNs considering leaving in the next year [Internet]. Nursing in Practice. 2024. Available from: https://www.nursinginpractice.com/latest-news/exclusive-28-of-gpns-considering-leaving-in-the-next-year/?contact_id=119984762&dm_t=0

5. Clifford J., Hitchinson, E, Hunter, H. Leading the Way: The role and value of nurses in general practice in England (Phase Three). Sonnet Advisory & Impact. 2024. Available from: https://sonnetimpact.co.uk/reports/the-role-and-value-of-gp-nurses-in-england/

6. Strategic Workforce Plan for Primary Care 2024/5-2029/30 https://heiw.nhs.wales/workforce/strategic-workforce-plan-for-primary-care/swppcsummary2024/#:~:text=Increased%20time%20for%20teaching%2C%20training,and%20retention%20in%20primary%20care.

7. Ford M. Future of primary care in Wales “at risk” amid ageing GPN workforce [Internet]. Nursing in Practice. 2024b. Available from: https://www.nursinginpractice.com/latest-news/future-of-primary-care-in-wales-at-risk-amid-ageing-gpn-workforce/

8. Halcomb Elizabeth J. and Ashley, Christine, “Australian primary health care nurses most and least satisfying aspects of work”. 2017. Faculty of Science, Medicine and Health - Papers: part A. 4760. https://ro.uow.edu.au/smhpapers/4760

9. Tort-Nasarre, G., Vidal-Alaball, J., Pedrosa, M., Vázquez Abanades,L., Forcada Arcarons A., and Rosanas JD. (2023). Factors associated with the attraction and retention of family and community medicine and nursing residents in rural settings: a qualitative study. BMC Medical Education, 2023 Sep 13(1). https://bmcmededuc.biomedcentral.com/articles/10.1186/s12909-023-04650-1

10. Anderson H, Scantlebury A, Galdas P, Adamson J. The well-being of nurses working in general practice during the COVID-19 pandemic: A qualitative study (The GenCo Study). Journal of Advanced Nursing. 2023 Oct 30 https://onlinelibrary.wiley.com/doi/10.1111/jan.15919

11. Ashley C, James S, Williams A, Calma K, Mcinnes S, Mursa R, et al. The psychological well-being of primary healthcare nurses during COVID-19: A qualitative study. Journal of Advanced Nursing. 2021;77(9):3820–8. 10.1111/jan.14937

12. Halcomb, E., Fernandez, R., Ashley, C., McInnes, S., Stephen, C., Calma, K., Mursa, R., Williams, A., & James, S.. The impact of COVID-19 on primary health care delivery in Australia. Journal of Advanced Nursing. 2022. 78(5), 1327– 1336. 10.1111/jan.15046

13. Mizumoto J, Mitsuyama T, Kumagaya S, Eto M, Izumiya M, Horita S. Primary care nurses during the coronavirus disaster and their struggle: Qualitative research. Journal of General and Family Medicine. 2022 Jul;23(5):343–50. 10.1002/jgf2.566

14. Russell A, de Wildt G, Grut M, Greenfield S, Clarke J. What can general practice learn from primary care nurses’ and healthcare assistants’ experiences of the COVID-19 pandemic? A qualitative study. BMJ Open. 2022 Mar;12(3):e055955. 10.1136/bmjopen-2021-055955

15. Bhardwa, S. Serious lack of practice nurses in leadership roles finds QNI research. Independent Nurse. 2016. Available from: https://www.independentnurse.co.uk/news/serious-lack-of-practice-nurses-in-leadership-roles-finds-qni-research/114524

16. Queen’s Nursing Institute. General practice nurse survey analysis. 2020. Queen’s Nursing Institute International Community Nursing Observatory. https://qni.org.uk/wp-content/uploads/2021/11/General-Practice-Nurses-Survey-Analysis-2020.pdf

17. Topping A. Exploring the implementation of the nursing associate role in general practice. Primary Health Care. 2023 Dec 4;33(6). doi: 10.7748/phc.2023.e1817 Available from: https://journals.rcni.com/primary-health-care/evidence-and-practice/exploring-the-implementation-of-the-nursing-associate-role-in-general-practice-phc.2023.e1817/full

18. Leary A, Maxwell E, Myers R, Punshon G. Why are healthcare professionals leaving NHS roles? A secondary analysis of routinely collected data. Human Resources for Health. 2024 Sep 20;22(1).. doi: 10.1186/s12960-024-00951-8. https://human-resources-health.biomedcentral.com/articles/10.1186/s12960-024-00951-8

19. Halcomb E, Smyth E, McInnes S. Job satisfaction and career intentions of registered nurses in primary health care: an integrative review. BMC Family Practice. 2018 Aug 7;19(1). https://bmcprimcare.biomedcentral.com/articles/10.1186/s12875-018-0819-1

20. Rendle KA, Abramson CM, Garrett SB, Halley MC, Dohan D. Beyond exploratory: a tailored framework for designing and assessing qualitative health research. BMJ Open. 2019 Aug;9(8):e030123. https://bmjopen.bmj.com/content/9/8/e030123

21. Jefferson L, Heathcote C, Bloor K. General practitioner wellbeing during the COVID-19 pandemic: a qualitative interview study. medRxiv. 2022 Jan 28. 10.1101/2022.01.26.22269874

22. Patton MQ. Qualitative research & evaluation methods: integrating theory and practice. Thousand Oaks: Sage Publications, Inc; 2015.

23. Braun V, Clarke V. To saturate or not to saturate? Questioning data saturation as a useful concept for thematic analysis and sample-size rationales. Qualitative Research in Sport, Exercise and Health. 2019 Dec 26;13(2):1–16.

24. Pope C, Mays N. Qualitative Research in Health Care. 4th ed. S.L.: Wiley-Blackwell; 2020.

25. Baker S, Edwards R. How Many Qualitative Interviews Is enough? Expert Voices and Early Career Reflections on Sampling and Cases in Qualitative Research [Internet]. 2012. Available from: https://eprints.ncrm.ac.uk/id/eprint/2273/4/how_many_interviews.pdf

26. GOV.UK. Data Protection Act [Internet]. GOV.UK. www.gov.uk; 2018. Available from: https://www.gov.uk/data-protection

27. Consolidated criteria for reporting qualitative research (COREQ): a 32-item checklist for interviews and focus groups | The EQUATOR Network [Internet]. Equator-network.org. 2015. Available from: https://www.equator-network.org/reporting-guidelines/coreq/

28. Smith B. Generalizability in qualitative research: misunderstandings, opportunities and recommendations for the sport and exercise sciences. Qualitative Research in Sport, Exercise and Health. 2018 Oct 23;10(1):137–49. DOI: 10.1080/2159676X.2017.139322. 10.1080/2159676X.2017.1393221

29. Holloway, I. (2008). A-Z of qualitative research in healthcare (2nd ed.). Wiley-Blackwell.

30. Willis, P. Raising the Bar. Shape of Caring: A Review of the Future Education and Training of Registered Nurses and Care Assistants. NHS England | Workforce, training and education. 2015. http://hee.nhs.uk/sites/default/files/documents/2348-Shape-of-caring-review-FINAL.pdf (Last accessed: 14 November 2024)

31. Nursing and Midwifery Council We Regulate Nursing Associates. 2023. http://www.nmc.org.uk/about-us/our-role/who-we-regulate/nursing-associates (Last accessed: 14 November 2024.)

32. NMC The NMC register 1 April 2022– 31 March 2023. Nursing and Midwifery Council. 2023. https://www.nmc.org.uk/globalassets/sitedocuments/data-reports/may-2023/0110a-annual-data-report-full-uk-web.pdf [last accessed 14th November 2024]

33. Peate I. British Journal of Nursing - The registered nursing associate: an overview [Internet]. British Journal of Nursing. 2023. Available from: https://www.britishjournalofnursing.com/content/nursing-associates/the-registered-nursing-associate-an-overview/

34. Health Education England (2021) Primary Care and General Practice Nursing Career and Core Capabilities Framework. Skills for Health. 2021 https://www.skillsforhealth.org.uk/resources/primary-care-general-practice-nursing-career-core-capabilities-framework/

35. Royal College of Nursing. General Practice Nursing staff: make sure you get your pay rise. The Royal College of Nursing. 2023. https://www.rcn.org.uk/news-and-events/news/uk-general-practice-nursing-staff-make-sure-you-get-your-pay-rise-210723#:~:text=The%20RCN%20is%20clear%20that%20salaried%20GP%20nursing,soon%20as%20possible%20and%20backdated%20to%20April%202023

36. Efendi F, Kurniati A, Bushy A, Gunawan J. Concept analysis of nurse retention. Nurs HealthSci. 2019;21:422–427. 10.1111/nhs.12629

37. Evans, N. Are nurse apprenticeship and associate schemes fit for purpose? Nursing standard, London: RCNi 2024-07, Vol.39 (7), p.29–31 https://rcni.com/nursing-standard/newsroom/analysis/are-nurse-apprenticeship-and-associate-schemes-fit-for-purpose-206536

38. Leary A, Punshon G, Hack A, Bushe D, Oldman C. The Impact of the Introduction of the Additional Roles Reimbursement Scheme on the General Practice Nursing Workforce in England. Journal of Primary Care & Community Health. 2024 Jan 1;1. doi: 10.1177/21501319241298759 https://journals.sagepub.com/doi/10.1177/21501319241298759

39. Holland PJ, Tham TL, Gill FJ. What nurses and midwives want: Findings from the national survey on workplace climate and well-being. International Journal of Nursing Practice. 2018 Feb 26;24(3):e12630. https://onlinelibrary.wiley.com/doi/10.1111/ijn.12630

40. Ford, M. Practice Nurses ‘diminished and belittled’ says BMA committee chair. Nursing in Practice 2024c file://hsci.york.ac.uk/HSCIYTU/Project%20--%20GenRet/Literature/Practice%20nurses%20%E2%80%98diminished%20and%20belittled%E2%80%99,%20says%20BMA%20committee%20chair%20_%20Nursing%20in%20Practice.html

41. Bovensiepen J, Pelkmans M. Dynamics of wilful blindness: An introduction. Critique of Anthropology. 2020 Oct 12;40(4):387–402.

42. Kim Y, Lee E, Lee H. Association between workplace bullying and burnout, professional quality of life, and turnover intention among clinical nurses. PLoS ONE. 2019.14(12): e0226506. 10.1371/journal. pone.0226506

43. Mawuena, EK., Mannion, R., Adu-Aryee, N.A., Adzei, F.A., Amoakwa, E.K., Twumasi, E. Professional disrespect between doctors and nurses: implications for voicing concerns about threats to patient safety. 2024. Vol 38. Iss 7. ISSN: 1477-7266 https://www.emerald.com/insight/content/doi/10.1108/jhom-06-2023-0167/full/html

44. Anderson M. Speaking up: the challenges and changes needed in primary care [Internet]. Nursing in Practice. 2024. Available from: https://www.nursinginpractice.com/analysis/speaking-up-the-challenges-and-changes-needed-in-primary-care/

45. Hult M, Ring M, Heta Siranko, Kangasniemi M. Decent and Precarious Work Among Nursing and Care Workers: A Mixed-Method Systematic Review. Journal of Advanced Nursing. 2024 Oct 25. 10.1111/jan.16572

46. Vives A, Amable M, Ferrer M, Moncada S, Llorens C, Muntaner C, et al. The Employment Precariousness Scale (EPRES): psychometric properties of a new tool for epidemiological studies among waged and salaried workers. Occupational and Environmental Medicine. 2010 Jun 24;67(8):548–55. 10.1136/oem.2009.048967.

47. Ejebu, O.Z., Philippou, J., Turnbull, J, et al., Coming and going: A narrative review exploring the push-pull factors during nurses’ careers, International Journal of Nursing Studies [Internet]. 2024 Sep 13 [cited 2024 Sep 28];160:104908–8. (2024) 10.1016/j.ijnurstu.2024.104908

48. Jones K, Wilson-Keates B, Melrose S. Using social media to recruit research participants: a literature review. Nurse Researcher. 2023 Dec 18;32(1). Available from: https://pubmed.ncbi.nlm.nih.gov/38105712/ 10.7748/nr.2023.e1859

