## Additional Table 1 for "Exploring the relationship between cultural and structural workforce issues and retention of nurses in general practice (GenRet): A qualitative interview study"

**Supplementary Table 1: All nursing roles in general practice by gender Dec 2023**

Data from NHS England 2024. General Practice Workforce Headcount and FTE Time series. Staff groups by job role and gender <https://app.powerbi.com/view?r=eyJrIjoiYTM4ZTA3NGItMTM2Mi00NzAwLWEyY2QtNDgyZDkxOTk3MmFlIiwidCI6IjUwZjYwNzFmLWJiZmUtNDAxYS04ODAzLTY3Mzc0OGU2MjllMiIsImMiOjh9>

| **Job role** | **Total** | **Female**  **(% of total)** | **Male**  **(% of total)** | **Other/unknown**  **(% of total)** |
| --- | --- | --- | --- | --- |
| **General Practice Nurses** | Headcount: 16,116  FTE: 11,368 | Headcount: 15,140  FTE: 10,858  (95.5%) | Headcount: 232  FTE: 190  (1.7%) | Headcount: 478  FTE: 319  (2.8%) |
| **Extended Role Practice Nurses** | Headcount: 769  FTE: 551 | Headcount: 722  FTE:514  (93.3%) | Headcount: 29  FTE: 24  (4.4%) | Headcount: 18  FTE: 13  (2.4%) |
| **Nurse specialists** | Headcount: 727  FTE: 484 | Headcount: 662  FTE: 435  (91.1%) | Headcount: 44  FTE: 34  (6.1%) | Headcount: 21`  FTE: 14  (1.9%) |
| **Advanced Nurse Practitioners** | Headcount: 5226  FTE:4069 | Headcount: 4666  FTE:3599  (88.5%) | Headcount: 428  FTE: 369  (9.1%) | Headcount: 134  FTE: 101  (3.3%) |
| **Nurse Partners** | Headcount: 74  FTE: 64 | Headcount: 66  FTE: 56  (87.5%) | Headcount: 6  FTE: 6  (9.4%) | Headcount: 2  FTE: 2  (3.1%) |
| **Other Nurses** | Headcount: 300  FTE: 221 | Headcount: 276  FTE:201  (91%) | Headcount: 18  FTE: 15  (6.8%) | Headcount: 6  FTE:5  (2.3%) |
| **Nursing Associates** | Headcount: 513  FTE: 420 | Headcount: 484  FTE: 397  (95%) | Headcount: 23  FTE:18  (4.3%) | Headcount: 6  FTE: 5  (1.2%) |
| **Trainee Nursing Associates** | Headcount: 228  FTE:216 | Headcount: 219  FTE: 201  (93%) | Headcount: 7  FTE: 7  (3.3%) | Headcount: 2  FTE: 2  (1%) |
| **Health Care Assistants** | Headcount: 9774  FTE: 7058 | Headcount: 9322  FTE: 6711  (95.1%) | Headcount: 321  FTE: 249  (3.5%) | Headcount: 135  FTE: 98  (1.4%) |

**Supplementary Table 2: All nursing roles in general practice by age (FTE) Dec 2023**

Data from NHS England 2024. General Practice Workforce FTE Time series. Staff groups by job role and age band <https://app.powerbi.com/view?r=eyJrIjoiYTM4ZTA3NGItMTM2Mi00NzAwLWEyY2QtNDgyZDkxOTk3MmFlIiwidCI6IjUwZjYwNzFmLWJiZmUtNDAxYS04ODAzLTY3Mzc0OGU2MjllMiIsImMiOjh9>

| **Job role** | **Age** | | | | | | | | | | |
| --- | --- | --- | --- | --- | --- | --- | --- | --- | --- | --- | --- |
|  | **<30**  **(% of total)** | **30-34**  **(% of total)** | **35-39**  **(% of total)** | **40-44**  **(% of total)** | **45-49**  **(% of total)** | **50-54**  **(% of total)** | **55-59**  **(% of total)** | **60-64**  **(% of total)** | **≥65**  **(% of total)** | **Unknown**  **(% of total)** | **Total** |
| **General Practice Nurses** | 915  (8.1%) | 1117  (9.8%) | 1231  (10.9%) | 1245  (11%) | 1311  (11.5%) | 1569  (13.8%) | 1785  (15.7%) | 1335  (11.7%) | 513  (4.5%) | 345  (3%) | 11366 |
| **Extended Role Practice Nurses** | 8  (1.5%) | 30  (5.5%) | 43  (7.8%) | 65  (11.8%) | 74  (13.5%) | 112  (20.4%) | 105  (19.1%) | 75  (13.6%) | 24  (4.4%) | 14  (2.5%) | 550 |
| **Nurse specialists** | 10  (2.1%) | 29  (6%) | 35  (7.3%) | 52  (10.7%) | 61  (12.6%) | 83  (17.2%) | 104  921.5%) | 72  (15%) | 27  (5.6%) | 10  (2%) | 583 |
| **Advanced Nurse Practitioners** | 40  (1%) | 161  (4%) | 310  (7.6%) | 507  (12.5%) | 642  (15.8%) | 856  (21%) | 825  (20.3%) | 461  (11.3%) | 156  (3.8) | 111  (2.7%) | 4069 |
| **Nurse Partners** | 2  (3.1%) | 1  (1.6%) | 3  (4.7%) | 8  (12.5%) | 13  (20.3%) | 11  (17.2%) | 9  (14.1%) | 11  (17.2%) | 4  (6.3%) | 2  (3.1%) | 64 |
| **Other Nurses** | 26  (11.8%) | 24  (11%) | 25  (11.4%) | 42  (19.1%) | 34  (15.5%) | 31  (14.1%) | 18  (8.9%) | 9  (4.1%) | 2  (0.9%) | 9  (4.1%) | 220 |
| **Nursing Associates** | 85  (20.3%) | 50  (11.9%) | 74  (17.4%) | 62  (14.8%) | 49  (11.7%) | 45  (10.7%) | 34  (8.1%) | 12  (2.9%) | 2  (0.5%) | 6  (1.4%) | 419 |
| **Trainee Nursing Associates** | 84  (40%) | 35  (16%) | 38  (18%) | 23  (11%) | 14  (6.7%) | 9  (4.3%) | 4  (1.9%) | 0  (0%) | 1  (0.5%) | 2  (1%) | 210 |
| **Health Care Assistants** | 740  (10.5%) | 641  (9.1%) | 691  (9.8%) | 756  (10.7%) | 798  (11.3%) | 1019  (14.4%) | 1149  (16.3%) | 839  (11.9%) | 321  4.5%) | 103  (1.5%) | 7057 |

**Table 3: All nurses working in general practice by ethnicity (Dec 2023)**

Data from NHS England 2024. General Practice Workforce. Staff group headcount by ethnicity

<https://app.powerbi.com/view?r=eyJrIjoiYTM4ZTA3NGItMTM2Mi00NzAwLWEyY2QtNDgyZDkxOTk3MmFlIiwidCI6IjUwZjYwNzFmLWJiZmUtNDAxYS04ODA>

| **Ethnicity** | **Headcount** | **% of total** |
| --- | --- | --- |
| **White** | 18,182 | 77.3 |
| **Asian/Asian British** | 850 | 3.6 |
| **Black/African/Caribbean/Black British** | 779 | 3.3 |
| **Other ethnic group** | 235 | 1 |
| **Mixed/multiple ethnic groups** | 196 | 0.83 |
| **Not recorded** | 3268 | 13.9 |
